# Associations between genetic HPV 16 diversity and cervical cancer prognosis

**DOI:** 10.1101/2024.08.02.24311429

**Authors:** Patrícia Patury, Fábio B. Russomano, Luiz F.L. Martins, Miguel Angelo Martins Moreira, Raquel B.M. Carvalho, Nádia R. C. Kappaun, Liz Maria de Almeida

## Abstract

**Introduction:** Cervical cancer (CC) arises as a result of chronic and persistent female genitalia infection by different oncogenic human papillomaviruses (HPV). The incidence of this disease is still high in developing countries such as Brazil, where the diagnosis is often made in advanced stages. HPV 16 is the most common type of CC worldwide. Studies concerning the association of different HPV 16 lineages with overall and disease-free CC survival rates can contribute to further understanding the behavior of different HPV 16 lineages concerning the prognosis of CC cases.

**Objective:** Assess the CC prognosis of patients treated in a Brazilian institution with regard to HPV16 strains.

**Methods:** Data were obtained from a prospective cohort of 334 CC patients recruited between July 2011 and March 2014 and treated at the Brazilian National Cancer Institute (INCA), in Rio de Janeiro, Brazil. HPV 16 lineages were identified in tumor tissue samples. Genetic HPV 16 diversity comprised 218 cases of lineage A, 10 of lineage B, 10 of lineage C and 96 of lineage D. In addition to HPV 16 lineages, age, histopathological type, staging, and treatment completion were evaluated regarding CC prognosis.

**Results:** Median patient age was 48 years old. The most common histopathological type was squamous cell carcinoma (82.3%), followed by adenocarcinoma. Locally advanced disease staging was the most frequently detected, represented by similar stage II and III percentages (36.2% and 37.7%), followed by initial stage I (19.2%) and stage IV presenting distant disease (6.9%). Only 187 patients completed CC treatment. Age, histological type, staging, and treatment completion were associated with a higher risk of death, which was not observed for the HPV 16 lineage variable. With regard to age, each one year of life increase led to about a 1% increase in risk of death. Other histopathological types (poorly differentiated carcinoma, adenosquamous, neuroendocrine and sarcoma) were associated with a higher risk of death compared to adenocarcinoma. Squamous cell carcinoma also represented a higher risk of death compared to adenocarcinoma, albeit non-statistically significant. Patients diagnosed in advanced stages exhibited a higher risk of death, and those who did not complete treatment exhibited an over 2-fold increased risk of death.

**Conclusion:** This study found no associations between HPV 16 lineages A, B, C and D and CC prognosis.

## Introduction

Cervical cancer (CC) is still a serious public health problem, especially in less developed countries, despite being considered a preventable disease through vaccination, screening, and treatment of precursor lesions. In countries with an opportunistic screening system, such as Brazil, or in those without public screening programs, CC diagnoses are often made when women already present advanced symptoms, compromising survival and quality of life [1].

Cervical cancer is the third most common cancer in Brazil among women excluding non-melanoma skin cancer. The number of new CC cases expected in Brazil is of 17,010 for each year of the 2023-2025 triennium, with a mortality rate of 6,627 deaths noted for 2021 [2, 3].

Different CC evolution processes in women with similar prognostic factors have not yet been fully clarified. Age, staging at diagnosis, lymphovascular space invasion, histopathological type and the presence of anemia are classically associated to CC prognoses [1, 4]. The prognostic value of oncogenic human papillomaviruses (HPV) DNA detection in tumors has not yet been established and studies with this objective have reported conflicting results. Some authors postulate that the presence of oncogenic HPV may be a useful CC prognosis marker prior to treatment, as this makes the precursor lesion progression more aggressive [5, 6]. Others, however, state that different types of oncogenic HPV have no prognostic value in CC cases [7, 8].

HPV 16 is the HPV most commonly associated with CC, and its four lineages were initially named according to the geographic region where they were most frequently identified, associated to local ethnicities [9, 10]. Since 2013, however, they have been termed A, B, C and D [11–14]. Each HPV 16 lineage (A, B, C and D) is subdivided into sublineages, as follows: A1-A3 (European and Asian lineages), A4 (Asian lineage); B1-B4 (African lineage 1); C1-C4 (African lineage 2); D1-D3 (North American and Asian-American lineages) [9, 10].

Some evidence suggests that different HPV16 lineages may present different pathogenicities, and some of them are associated with a greater risk of developing CC than others [15]. Certain authors have demonstrated that lineages B, C and D play a more important role in the progression of precursor cervix lesions in the invasive CC when compared to lineage A [9, 16, 17]. Other studies, however, have suggested that the D lineage, in particular, is highly associated with viral infection persistence and progression to CC [18, 19, 20].

Regarding the prognostic value of different HPV16 lineages and sublineages in CC cases, Tornesello et al. [21] suggested that the AA (Asian-American) lineage was associated to more aggressive CC behavior. Another study evaluated 301 *in situ* cases and 727 CC cases and reported a higher frequency of HPV 16 D3 and A4 in CC cases and lower disease-free survival rates in women infected with the HPV 16 B lineage compared to lineages A, C and D [22]. In another assessment, Zuna et al. [23] reported more advanced stages (II-IV) and lower survival rates in women infected with the European HPV 16 lineage when assessing 155 CC patients.

In view of these contradictions and the possibility that differences may be associated with population characteristics or influenced by treatment strategies, this study aimed to assess whether the HPV 16 lineages present in CC cases treated at the Brazilian National Cancer Institute (INCA) in Rio de Janeiro, Brazil, influence CC prognosis regarding overall survival and progression-free survival.

## Methods

### Patients

This prospective cohort study took place at a specialized gynecological oncology center in Brazil from July 2011 to March 2014. It selected women aged 18 years or older, diagnosed with FIGO-2009 IB1 or higher cervical cancer (CC), and excluded patients who had undergone previous cancer treatment [17, 24]. Over the course of a five-year duration, all participants were monitored, with their clinical data collected until January of 2022. All participants provided written informed consent prior to participation. Ethical approval for the study was provided by the Human Research Ethics Committee at the oncology center.

Nine hundred and sixty-eight women with a histopathological diagnosis of CC were included. All were 18 years old or older and had not undergone previous oncological CC treatment. The HPV types and lineages in positive HPV16 cases were identified by tumor tissue biopsies obtained at the first consultation for 594 (61.4%) patients. New samples were not obtained for the remaining cases due to technical difficulties, poor clinical patient conditions or refusal to undergo the procedure. The samples were immediately stored in RNA-Later at -80°C and deposited in the National Cancer Institute Tumor Bank.

### DNA isolation and identification of HPV16 genotypes and lineages

Total DNA was isolated from the obtained biopsies using the QIAamp DNA Mini Kit (Qiagen, Hilden, Germany) and eluted in 200 microliters (µL) of AE buffer and stored at – 80°C. After isolation, the presence of viral DNA was detected by polymerase chain reactions (PCR) with a set of PGMY09/11 primers [25]. When DNA was not detected by this technique, a nested PCR was performed employing the GP5p/GP6p primers [17, 26].

The amplified PCR products were purified using the GFX PCR and DNA Band Purification kit (GE Healthcare), sequenced with the Big Dye Terminator v3.1 Cycle Sequencing Kit (Applied Biosystems, Foster City, CA) and analyzed using an ABI Prism. 3130XL Genetic Analyzer (Applied Biosystems). All sequences were edited and analyzed using the 4Peaks Software (Nucleobytes, Amsterdam, Netherlands). HPV genotypes were identified using the Basic Local Alignment Search Tool (BLASTn, http://blast.ncbi.nlm.nih.gov/) [17]. The biopsies presenting HPV16 genotypes (n=392) were submitted to PCR for amplification of the viral genomic regions *E6* and *LCR*, followed by DNA sequencing. This allowed the identification of different haplotypes, and HPV16 lineages and were reported previously in Vidal et al. (2016) [27]. HPV16 lineages were identified in 334 samples Fig 1.

**Fig 1.**
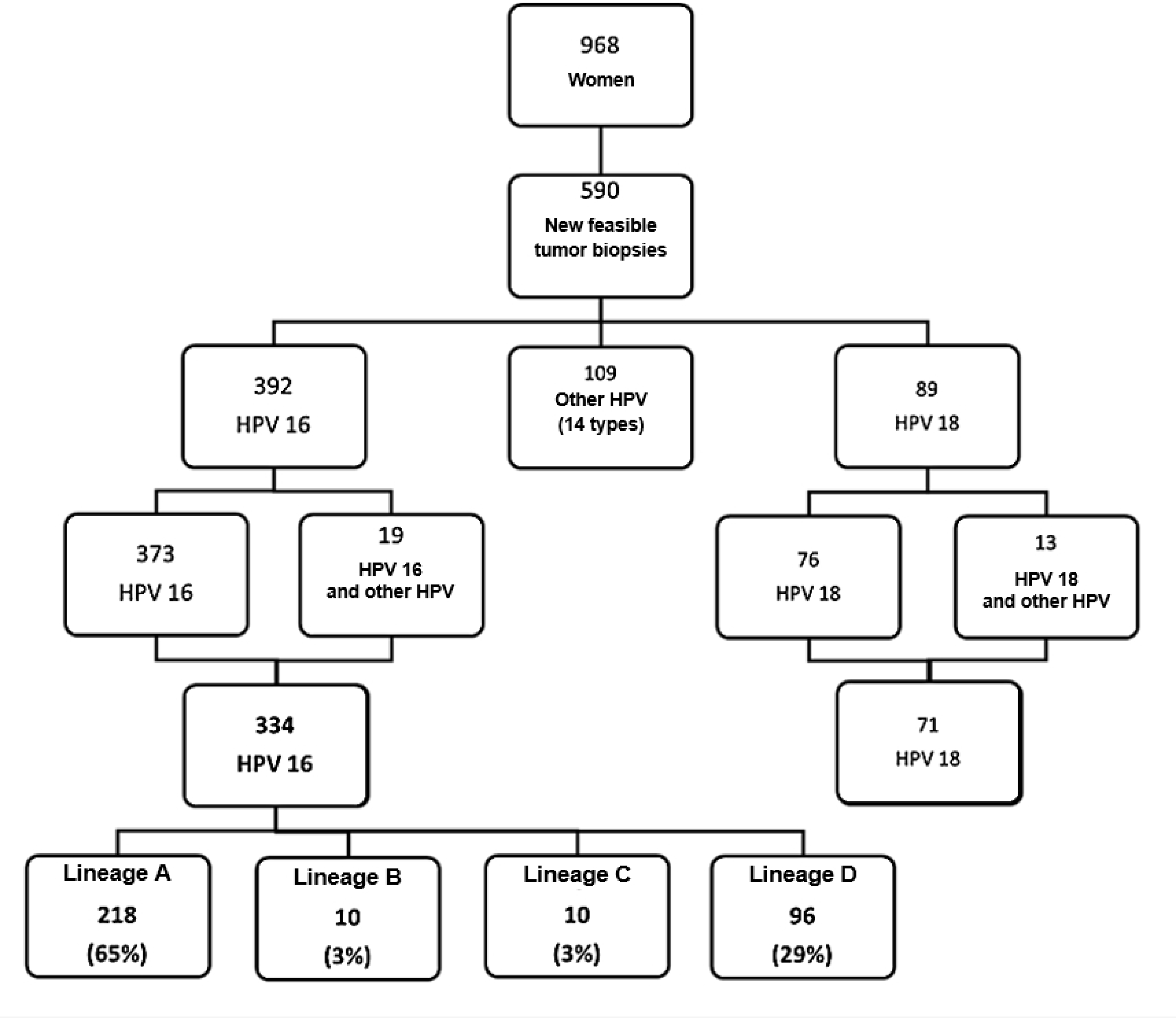
Flowchart depicting patient inclusion and types of detected HPV. (INCA, 2011-2014).

### Socioeconomic data

A questionnaire was applied to obtain educational, environmental, and socioeconomic characteristics of all enrolled patients. Clinical information concerning treatment and follow-up regarding patients infected with HPV16 was obtained from both physical and electronic records.

### Statistical analyses

Crude and adjusted prevalence ratios and their respective 95% confidence intervals (95% CI) were calculated by means of the Poisson regression model with variations to determine associations between HPV 16 lineages and other variables [28, 29]. A multinominal logistic regression model was applied concerning histological type, due to three histological categories. Models presenting p values < 0.05 were considered significant.

Kaplan-Meyer curves were used in a univariate and stratified manner for confounding variables for the survival analyses. A semiparametric proportional risk model (Cox model) was used for the multivariate analyses. Two Schoenfeld residuals were analyzed to verify model validity, and associations were considered significant when p < 0.05.

Overall survival rates were calculated setting the survival time as the interval between the data of histopathological revision of the biopsy for diagnosis confirmation and the date of death or last follow-up. Recurrence or recurrence-free survival times were defined in months, considering the interval between the date of the end of the treatment and the date of the recurrence or the date of the last consultation with no CC signs. Patients presenting disease progression throughout the treatment or at treatment end were not included in the disease-free survival analysis.

All procedures were approved by the INCA Ethics and Research Committee (number 156/10, on 02/25/2011) and a specific amendment for this study (CAAE: 53398416.0.0000.5274, on 03/01/2016) and all the patients signed a consent form.

## Results

Patient and tumor characteristics are depicted in Table 1. Median patient age was 48 years old. Most women carried lineages A (65.3%) and D (28.7%), while lineage B was detected in 3% of all cases and lineage C, in another 3%.

**Table 1.** Description of the study population (n = 334).

| Characteristics | n | % |
| --- | --- | --- |
| **Sociodemographic** |  |  |
| **Age (years)** |  |  |
| Up to 39 | 96 | 28.7 |
| 40 to 49 | 93 | 27.8 |
| 50 to 64 | 106 | 31.7 |
| 65 and over | 39 | 11.7 |
| **Years of study** |  |  |
| None | 19 | 5.7 |
| 1 to 3 | 70 | 21.0 |
| 4 to 7 | 108 | 32.3 |
| 8 to 10 | 82 | 24.6 |
| 11 and over | 55 | 16.5 |
| **Marital status** |  |  |
| Single | 28 | 8.4 |
| Married | 184 | 55.1 |
| Divorced | 71 | 21.3 |
| Widow | 51 | 15.3 |
| **Skin color** |  |  |
| White | 108 | 32.3 |
| Brown | 178 | 53.3 |
| Black | 46 | 13.8 |
| Yellow | 2 | 0.6 |
| ***HPV 16 genotyping (detected lineages)*** |  |  |
| A | 218 | 65.3 |
| B | 10 | 3.0 |
| C | 10 | 3.0 |
| D | 96 | 28.7 |
| ***Tumor and treatment features*** |  |  |
| **Histological Type** |  |  |
| Squamous cell carcinoma | 275 | 82.3 |
| Adenocarcinoma | 43 | 12.9 |
| Others<sup>1</sup> | 16 | 4.8 |
| **Staging** |  |  |
| I | 64 | 19.2 |
| II | 121 | 36.2 |
| III | 126 | 37.7 |
| IV | 23 | 6.9 |
| **Treatment completion** |  |  |
| Yes | 187 | 56.0 |
| No | 147 | 44.0 |
| **Outcome** |  |  |
| **Persistence** |  |  |
| Yes | 105 | 31.4 |
| No | 229 | 68.6 |
| No data |  |  |
| **Relapse (of the 229 patients with no disease persistence)** |  |  |
| Yes | 80 | 34.9 |
| No | 149 | 65.1 |
| **Death** |  |  |
| Yes | 192 | 57.5 |
| No | 142 | 42.5 |
| **Death due to cervical cancer** |  |  |
| Yes | 174 | 52.1 |
| No | 18 | 5.4 |

(<sup>1</sup>) 6 G3 Carcinoma cases, 5 Adenosquamous cases, 4 Neuroendocrine cases and 1 Sarcoma case.

The most common histopathological type was squamous cell carcinoma (82.3%), followed by adenocarcinoma (12.9%). Stages II and III were the most common (36.2% and 37.7%), followed by stage I (19.2%) and IV (6.9%). Only 187 women completed treatment. Regarding treatment results, about 1/3 of the patients exhibited disease persistence (31.4%), most displayed disease remission and did not relapse (65%) and 192 died during the follow-up period Table 1.

### Overall Survival

The median follow-up time for the enrolled patients was of 35.9 months (1Q = 12.6 months; 2Q = 36.3 months and 3Q = 66 months), and the median overall survival time was of 40.3 months (CI_95%_: 29.9 – 49.8).

Kaplan-Meyer curves according to each HPV 16 lineage are displayed in Fig 2. The median overall survival was of 35.9 months among patients carrying lineage A and 45.9 months among those carrying lineage D. No estimated median was determined for the B/C group, as the survival curve was influenced by sample size. No statistically significant difference between the survival curves (p = 0.21; Log Rank test) was observed.

**Fig 2.**
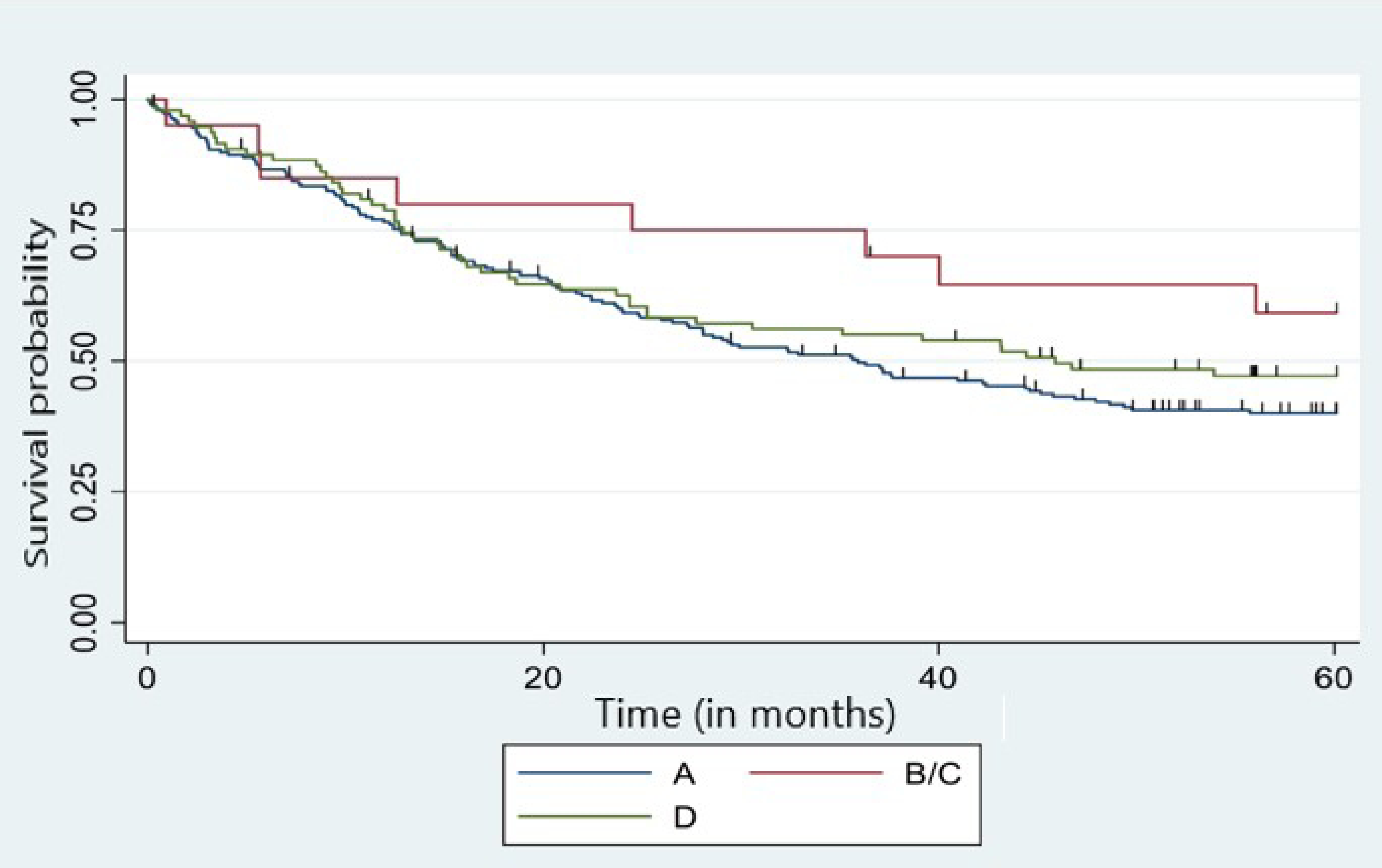
Five-year overall survival curve of cervical cancer patients since the beginning of the treatment according to HPV 16 lineage*. Lineage A = 218 cases, B = 10 cases, C = 10 cases, D = 96 cases. *HPV 16 lineages B and C were grouped due to their low frequency.

Table 2 indicates risk of patient death according to the study variables. Age, histological type, treatment completion and staging were associated with a higher risk of death, while HPV 16 lineage was not. Concerning age, about a 1% increase in the risk of death was noted for each one year of life increase.

**Table 2.** Overall risk of death in patients with cervical cancer (n = 334)

| Characteristic | n | Crude HR<br>(CI 95%) | P value |
| --- | --- | --- | --- |
| **HPV 16 lineage** |  |  |  |
| A | 218 | 1.18 (0.85 - 1.64) | 0.32 |
| B/C | 20 | 0.67 (0.32 - 1.41) | 0.29 |
| D | 96 | 1 | - |
| **Age (years)** |  | 1.01 (1.00 - 1.02) | 0.02 |
| **Histological type** |  |  |  |
| Squamous cell carcinoma | 275 | 1.51 (0.94 - 2.43) | 0.09 |
| Adenocarcinoma | 43 | 1 | - |
| Others | 16 | 2.47 (1.17 - 5.19) | 0.02 |
| **Staging** |  |  |  |
| I or II | 185 | 1 | - |
| III or IV | 149 | 2.69 (2.00 - 3.61) | < 0.001 |
| **Treatment completion** |  |  |  |
| Yes | 187 | 1 |  |
| No | 147 | 4.06 (3.00 - 5.48) | < 0.001 |

HR: hazard ratio.

Tumors characterized as other histopathological types (poorly differentiated carcinoma, adenosquamous, neuroendocrine and sarcoma) were associated to a higher risk of death compared to adenocarcinoma (HR: 2.47) (95% CI 1.17 - 5.19). Patients presenting squamous cell carcinomas also exhibited a higher risk of death than those presenting adenocarcimomas, although non-statistically significant Table 2.

Women diagnosed in advanced stages displayed a higher risk of death and those who did not complete treatment presented a more than two-fold increased risk of death (HR: 2.69 (2.00 - 3.61)) Table 2.

The staging, histological type and treatment completion data distribution suggest an association between intermediate variables and HPV 16 lineages, which was not observed for age. Considering the possibility that different HPV 16 lineages could be associated with variables that could lead to worse CC prognoses, prevalence ratios between HPV 16 lineages and other variables explaining worse prognosis were calculated Table 3. The association between HPV 16 lineages, staging and treatment completion reached borderline significance (p=0.07).

**Table 3.** Prevalence ratios between HPV 16 lineages and explanatory variables for worse prognosis.

| Characteristic | Advanced staging<sup>1</sup> |  | Histological type<sup>2</sup> |  | Did not finish treatment |  |
| --- | --- | --- | --- | --- | --- | --- |
|  | Prevalence ratio*<br>(CI 95%) | p value | Prevalence ratio*<br>(CI 95%) | p value | Prevalence ratio*<br>(CI 95%) | p value |
| HPV 16 lineage |  |  |  |  |  |  |
| A | 1.30 (0.97 - 1.74) | 0.07 | 0.70 (0.35 - 1.38) | 0.30 | 1.31 (0.98 - 1.75) | 0.07 |
| B/C | 0.82 (0.40 - 1.67) | 0.54 | 1.86 (0.52 - 6.73) | 0.34 | 0.80 (0.39 - 1.64) | 0.54 |
| D | 1 |  |  |  | 1 | - |

<sup>1</sup> – Staging III and IV
<sup>2</sup> - Histological type (Squamous cell carcinoma, Adenocarcinoma, others); using the multiple logistic regression model

### Disease-Free Survival

Up to 75% of all patients exhibited a disease-free timeframe lower than 23.9 months. The Kaplan-Meyer disease-free survival curves according to HPV 16 lineage are presented in Fig 3. The third survival quartile was of 23.5 months among patients carrying lineage A, 31.1 for patients carrying the B/C lineages and 24.9 months for those carrying lineage D. No estimated median was calculated for group B/C due to the number of patients carrying this lineage. No statistically significant difference between survival curves was noted when applying the log-rank test (p = 0.43).

**Fig 3.**
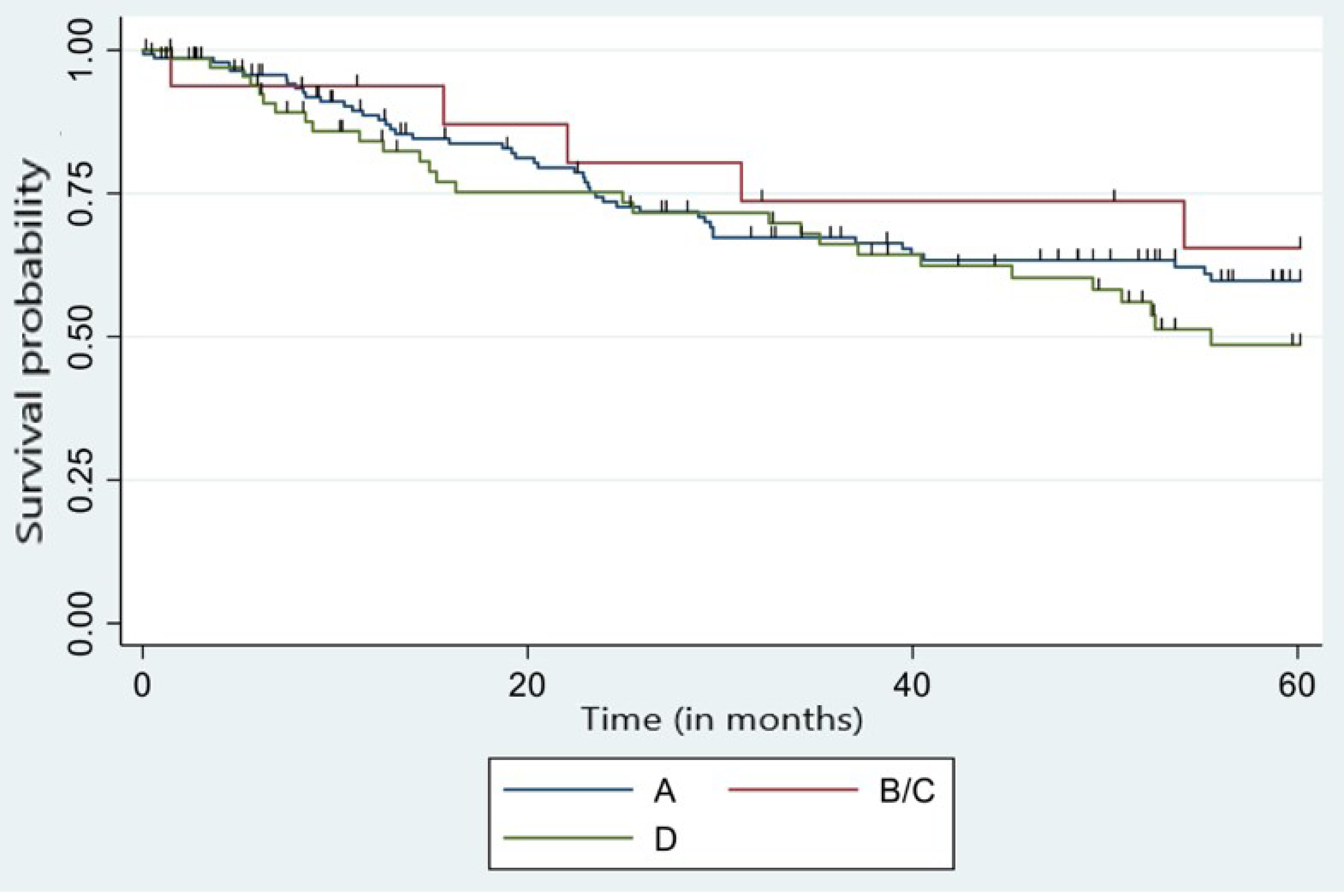
Disease-free survival curve in 5 years of patients with cervical cancer since the end of treatment according to HPV 16 lineage*. *Patients who did not exhibit persistent cervical cancer after treatment. *HPV 16 lineages B and C were grouped due to their low frequency.

Staging was the only variable significantly associated with the risk of CC recurrence, expressed by risk ratios. In this case, women presenting advanced staging exhibited a HR of 1.83 (95%CI 1.17 - 2 .87) compared to those presenting early staging Table 4.

**Table 4.** Risk of cervical cancer recurrence (n = 229*).

| Characteristic | n | Crude HR<sup>†</sup><br>(CI 95%) | P value |
| --- | --- | --- | --- |
| **HPV 16 lineage** |  |  |  |
| A | 218 | 0.77 (0.48 - 1.23) | 0.28 |
| B/C | 20 | 0.35 (0.13 - 1.00) | 0.3 |
| D | 96 | 1 | - |
| **Age (years)** |  | 1.00 (0.99 - 1.02) | 0.59 |
| **Histological type** |  |  |  |
| Squamous cell carcinoma | 275 | 0.70 (0.41 – 1.18) | 0.18 |
| Adenocarcinoma | 43 | 1 | - |
| Others | 16 | 0.24 (0.03 – 1.83) | 0.17 |
| **Staging** |  |  |  |
| I or II | 185 | 1 |  |
| III or IV | 149 | 1.83 (1.17 - 2.87) | 0.008 |
| **Treatment completion** |  |  |  |
| Yes | 187 | 1 | - |
| No | 147 | 1.14 (0.68 – 1.92) | 0.61 |

\* Patients who did not exhibit persistence post-treatment cervical cancer,
†HR = *hazard ratio*

## Discussion

This study aimed to assess the influence of HPV 16 strains on CC prognosis. In this sense, only age, histopathological type, disease staging, and treatment completion were associated with a higher risk of death.

No associations between HPV 16 lineages and CC prognosis were noted, although different prevalences of each were detected according to stage, with no confounding effect demonstrated between these variables Table 3. The low frequencies of lineages B and C may have contributed to the lack of statistically significant associations noted herein. The most frequently detected lineage was the A (Asian-European) lineage, in contrast to that reported by Tornesello et al. [30] (Asian-American), which was termed lineage D in this study. As suggested by Tornesello et al. [30], lineage D exhibited more aggressive behavior in CC cases [30].

Although no significant CC prognosis differences were detected in relation to HPV 16 lineages, other authors have noted this association. Zuna et al. [31], for example, reported lower survival rates in women carrying the European lineage. In our study, the better survival rates observed for patients carrying non-European lineages (B, C and D) were partially mediated by initial staging at diagnosis, suggesting that this lineage is associated to less aggressive tumor behavior compared to the European lineage. In another study, Rader et al. [32] reported lower disease-free survival rates in patients carrying the HPV16 B lineage.

The median age of CC patients in the present study was 48 years old, and a 1% increase in the risk of death was noted with each increasing year. This finding is in agreement with studies that have demonstrated age as an independent CC prognostic factor [33–38].

Squamous cell carcinoma was the most common histopathological type detected in the present study, representing 82.3% of all cases Table 1, in line with other literature reports [1]. The “others” histopathological category defined herein (poorly differentiated carcinoma, adenosquamous, neuroendocrine and sarcoma) presented a higher risk of death in relation to adenocarcinoma Table 2. These tumors exhibit a more aggressive clinical evolution and are generally diagnosed at more advanced stages [39, 40].

Contrary to other authors, who report worse prognosis for adenocarcinoma cases [41–44], a higher risk of death for patients presenting squamous cell carcinoma was observed herein, albeit not statistically significant. This may be due to the fact that most adenocarcinoma cases were detected in stage I.

Staging was the only significant variable regarding CC recurrence risk, in agreement with other authors [1, 45]. Furthermore, patients exhibited a 4-fold increase concerning risk of death when they did not complete the treatment. Most women unable to complete treatment presented advanced CC stages and exhibited associated comorbidities, such as anemia and kidney failure, making it impossible to complete initial therapeutic plans. Other authors have associated treatment completion and other variables to age, reporting that CC patients aged 70 or over exhibit a higher rate of less aggressive treatment or were unable to receive treatment at all, although they do not mention why no treatment or incomplete treatment took place [33].

An unequal incomplete treatment distribution according to HPV 16 lineage Table 4 was noted in the present study, where just over half of the patients carrying lineages A and D did not complete the treatment, while 80% of those carrying the B/C lineages did so. This may have been influenced by lineage staging distribution.

Although no significant association between HPV 16 strains and CC prognosis was observed, few studies have assessed this issue to date. This association may not have been detected in the present study due to the limited sample size for certain HPV 16 lineages. Studies encompassing larger sample sizes will be able elucidate this issue and guide new treatment protocols and better information for patients if associations are demonstrated.

## Conclusion

No statistically significant associations between HPV 16 lineages and CC prognosis were observed herein. Age, poorly differentiated carcinoma, adenosquamous, neuroendocrine and sarcoma histopathological types, advanced staging and incomplete treatment were associated with worse CC prognoses. Considering the sample size limitations of some of HPV16 strains in this study, their influence on CC prognosis cannot be disregarded, and further studies in this regard may contribute to elucidate this issue.

## Data Availability

Our data sets will be available upon request. Data are available from the Ethics Committee (contact via +552132074550) for researchers who meet the criteria for access to confidential data.

## Acknowledgments

We thank all the women treated at INCA in Rio de Janeiro, for allowing this project to be carried out. We also thank the interdisciplinary teams that participated in the treatment and follow up of these women.

## References

1. Marth C, Landoni S, Mahner S, et al. Cervical Cancer: ESMO Clinical Practice Guidelines for Diagnosis, Treatment and Follow-Up. Ann Oncol [Internet]. 2017 [citado em: 23 de novembro de 2021]; 28(suppl_4):iv72-iv83. Erratum in: Ann Oncol. 2018 1;29 (Suppl 4): iv262. Disponível em: https://pubmed.ncbi.nlm.nih.gov/28881916/. Doi: 10.1093/annonc/mdx220. PMID: 28881916.

2. BRASIL. Ministério da Saúde. INCA - Instituto Nacional de Câncer José Alencar Gomes da Silva [Internet]. Estatísticas de câncer. [Publicado em 23 de junho de 2022, atualizado em 24 de novembro de 2022]. [citado em: 30 de novembro de 2022]. Disponível em: https://www.gov.br/inca/pt-br/assuntos/cancer/numeros.

3. BRASIL. Ministério da Saúde. INCA - Instituto Nacional de Câncer José Alencar Gomes da Silva. Detecção Precoce. [Internet]. 2022 [citado em: 30 de novembro de 2022]. Publicado em 2022. Disponível em: https://www.gov.br/inca/pt-br/assuntos/gestor-e-profissional-de-saude/controle-do-cancer-do-colo-do-utero/acoes/deteccao-precoce.

4. Koulis TA, Kornaga EN, Banerjee R, et al. Anemia, Leukocytosis and Thrombocytosis as Prognostic Factors in Patients with Cervical Cancer Treated with Radical Chemoradiotherapy: A Retrospective Cohort Study. Clin Transl Radiat Oncol [Internet]. 2017 [citado em: 09 de julho de 2022]; 4;51-56. Disponível em: https://pubmed.ncbi.nlm.nih.gov/29594208/. Doi: 10.1016/j.ctro.2017.05.001. PMID: 29594208; PMCID: PMC5833917.

5. Li Ping, Tan Yue, Zhu Li-Xia, et al. Prognostic Value of HPV DNA Status in Cervical Cancer Before Treatment: A Systematic Review and Meta-Analysis. Oncotarget [Internet]. 2017 [citado em: 04 de setembro de 2021]; 8(39):66352–66359. Disponível em: https://pubmed.ncbi.nlm.nih.gov/29029517/. Doi: 10.18632/oncotarget.18558. PMID: 29029517; PMCID: PMC5630417.

6. Chong GO, Lee YH, Soo Han H, et al. Prognostic Value of Pre-Treatment Human Papilloma Virus DNA Status in Cervical Cancer. Gynecologic Oncology [Internet]. 2017 [citado em: 02 de junho de 2022]; p97-102. Disponível em: https://pubmed.ncbi.nlm.nih.gov/29153540/. Doi: 10.1016/j.ygyno.2017.11.003. PMID: 29153540.

7. Ikenberg H, Sauerbrei W, Schottmüller U, et al. Human Papillomavirus Dna in Cervical Carcinoma—Correlation with Clinical Data and Influence on Prognosis. Int. J. Cancer [Internet]. 1994 [citado em: 05 de agosto de 2022]; 59 (3)322–326. Disponível em: https://europepmc.org/article/MED/7927936. DoiI: 10.1002/ijc.2910590306. PMID: 7927936.

8. Füle T, Csapó Z, Máthé M, et al. Prognostic Significance of High-Risk Hpv Status in Advanced Cervical Cancers and Pelvic Lymph Nodes. Gynecol Oncol [Internet]. 2006 [citado em: 09 de outubro de 2022]; 100 (3) 570–578. Epub 2005. Disponível em: https://pubmed.ncbi.nlm.nih.gov/16325245/. doi: 10.1016/j.ygyno.2005.09.019. PMID: 16325245.

9. Clifford GM, Tenet V, Georges D, et al. Human papillomavirus 16 sub- lineage dispersal and cervical cancer risk worldwide: whole viral genome sequences from 7116 HPV16- positive women. Papillomavirus Res [Internet]. 2019 [citado em: 16 de setembro de 2021]; 7:67-74. Disponível em: https://pubmed.ncbi.nlm.nih.gov/30738204/. Doi: 10.1016/j.pvr.2019.02.001. PMID: 30738204; PMCID: PMC6374642.

10. Mirabello L, Clarke MA, Nelson CW, et al. The Intersection of HPV Epidemiology, Genomics and Mechanistic Studies of HPV-Mediated Carcinogenesis. Viruses [Internet]. 2018 [citado em: 09 de outubro de 2022]; 10(2):80. Disponível em: https://pubmed.ncbi.nlm.nih.gov/29438321/. Doi: 10.3390/v10020080. PMID: 29438321; PMCID: PMC5850387.

11. Ho L, Chan S, Burk R, et al. The Genetic Drift of Human Papillomavirus Type 16 Is a Means of Reconstructing Prehistoric Viral Spread and the Movement of Ancient Human Populations. J Virol [Internet]. 1993 [citado em: 08 de abril de 2022]; 16:6413–6423. Disponível em: https://journals.asm.org/doi/10.1128/jvi.67.11.6413-6423.1993.

12. Ong CK, Chan SY, Campo MS, et al. Evolution of Human Papillomavirus Type 18: An Ancient Phylogenetic Root in Africa and Intratype Diversity Reflect Coevolution with Human Ethnic Groups. J Virol [Internet]. 1993 [citado em: 08 de abril de 2022]; 67(11):6424–6431. Disponível em: https://pubmed.ncbi.nlm.nih.gov/8411344/. Doi: 10.1128/JVI.67.11.6424-6431.1993. PMID: 8411344; PMCID: PMC238077.

13. Yamada T, Manos MM, Peto J, et al. Human Papillomavirus Type 16 Sequence Variation in Cervical Cancers: A Worldwide Perspective. J Virol [Internet]. 1997 [citado em: 03 de março de 2022]; 71(3):2463-72. Disponível em: https://pubmed.ncbi.nlm.nih.gov/9032384/. Doi: 10.1128/JVI.71.3.2463-2472.1997. PMID: 9032384; PMCID: PMC191357.71:2463–2472.

14. Burk RD, Terai M, Gravitt PE, et al. Distribution of human papillomavirus types 16 and 18 variants in squamous cell carcinomas and adenocarcinomas of the cervix. Cancer Res [Internet]. 2003 [citado em: 09 de abril de 2022]; 63(21):7215–20. Disponível em: https://pubmed.ncbi.nlm.nih.gov/14612516.

15. Margolis B, Cagle-Colon K, Chen L, et al. Prognostic Significance of Lymphovascular Space Invasion for Stage Ia1 and Ia2 Cervical Cancer. Int J Gynecol Cancer [Internet]. 2020 [citado em: 07 de março de 2022]; 30:735-743. Disponível em: https://pubmed.ncbi.nlm.nih.gov/32179697/. Doi: 10.1136/ijgc-2019-000849. PMID: 32179697.

16. Chen JL, Huang C-Y, Huang Y-S, et al. Differential clinical characteristics, treatment response and prognosis of locally advanced adenocarcinoma/adenosquamous carcinoma and squamous cell carcinoma of cervix treated with definitive radiotherapy. Acta Obstet Gynecol Scand [Internet]. 2014 [citado em: 01 de março de 2023]; 93(7):661-8. Disponível em: https://pubmed.ncbi.nlm.nih.gov/24666257/. Doi: 10.1111/aogs.12383. PMID: 24666257.

17. Vidal JPCB, Felix SP, Chaves CBP, et al. Genetic Diversity of HPV16 and HPV18 in Brazilian Patients with Invasive Cervical Cancer. J Med Virol [Internet]. 2016 [citado em: 14 de março de 2023]; 88(7):1279-87. Disponível em: https://pubmed.ncbi.nlm.nih.gov/26694554/. Doi: 10.1002/jmv.24458. PMID: 26694554.

18. Schiffman M, Rodriguez AC, Chen Z, et al. A population-based prospective study of carcinogenic human papillomavirus variant lineages, viral persistence, and cervical neoplasia. Cancer Res [Internet]. 2010 [citado em: 30 de abril de 2023]; 70(8):3159-69. Disponível em: https://pubmed.ncbi.nlm.nih.gov/20354192/. Doi: 10.1158/0008-5472.CAN-09-4179. PMID: 20354192; PMCID: PMC2855741.

19. Berumen J, Ordoñez RM, Lazcano E, et al. Asian-American variants of human papillomavirus 16 and risk for cervical cancer: a case-control study. J Natl Cancer Inst [Internet]. 2001 [citado em:02 de dezembro de 2022]; 5;93(17):1325-30. Disponível em: https://pubmed.ncbi.nlm.nih.gov/11535707. Doi: 10.1093/jnci/93.17.1325. PMID: 11535707.

20. Xi LF, Koutsky LA, Hildesheim A, et al. Risk for high-grade cervical intraepithelial neoplasia associated with variants of human papillomavirus types 16 and 18. Cancer Epidemiol Biomarkers Prev [Internet]. 2007 [citado em: 05 de dezembro de 2022]; 16(1):4-10. Disponível em: https://pubmed.ncbi.nlm.nih.gov/17220325/. Doi: 10.1158/1055-9965.EPI-06-0670. PMID: 17220325.

21. Tornesello ML, Losito S, Benincasa G, et al. Human papillomavirus (HPV) genotypes and HPV16 variants and risk of adenocarcinoma and squamous cell carcinoma of the cervix. Gynecol Oncol [Internet]. 2011 [citado em: 10 de dezembro de 2022]; 121(1):32-42. Disponível em: https://pubmed.ncbi.nlm.nih.gov/21211829/. Doi: 10.1016/j.ygyno.2010.12.005. PMID: 21211829.

22. Rader JS, Tsaih SW, Fullin D, et al. Genetic variations in human papillomavirus and cervical cancer outcomes. Int J Cancer [Internet]. 2019 [citado em: 03 de janeiro de 2023]; 144(9): 2206–2214. Disponível em: https://pubmed.ncbi.nlm.nih.gov/30515767/. Doi:10.1002/ijc.32038. PMID: 30515767; PMCID: PMC6450540.

23. Zuna RE, Tuller E, Wentzensen N, et al. HPV16 variant lineage, clinical stage, and survival in women with invasive cervical cancer. Infect Agents Cancer [Internet]. 2011 [citado em: 23 de abril de 2023]; 6:19. Disponível em: 10.1186/1750-9378-6-19.

24. de Almeida LM, Martins LFL, Pontes VB, et al. Human Papillomavirus Genotype Distribution among Cervical Cancer Patients prior to Brazilian National HPV Immunization Program. J Environ Public Health [Internet]. 2017 [citado em: 04 de dezembro de 2022]; v. 2017:1645074, p.9. Disponível em: https://pubmed.ncbi.nlm.nih.gov/28512474/. Doi: 10.1155/2017/1645074. PMID: 28512474; PMCID: PMC5420420.

25. Gravitt PE, Peyton CL, Alessi TQ, et al. Improved amplification of genital human papillomaviruses. J Clin Microbiol [Internet]. 2000 [citado em: 01 de fevereiro de 2023]; 38:357–361. Disponível em: https://pubmed.ncbi.nlm.nih.gov/10618116/. Doi: 10.1128/JCM.38.1.357-361.2000. PMID: 10618116; PMCID: PMC88724.

26. Fuessel Haws AL, He Q, Rady PL, et al. Nested PCR with the PGMY09/11 and GP5(+)/6(+) primer sets improves detection of HPV DNA in cervical samples. J Virol Methods [Internet]. 2004 [citado em: 13 de fevereiro de 2023]; 122:87–93. Disponível em: https://pubmed.ncbi.nlm.nih.gov/15488625/. doi: 10.1016/j.jviromet.2004.08.007. PMID: 15488625.

27. Vidal JPCB, Felix SP, Chaves CBP, et al. Genetic Diversity of HPV16 and HPV18 in Brazilian Patients with Invasive Cervical Cancer. J Med Virol [Internet]. 2016 [citado em: 14 de março de 2023]; 88(7):1279-87. Disponível em: https://pubmed.ncbi.nlm.nih.gov/26694554/. Doi: 10.1002/jmv.24458. PMID: 26694554.

28. Zocchetti C, Consonni D, Bertazzi PA. Relationship between prevalence rate ratios and odds ratios in cross-sectional studies. Int J Epidemiol [Internet]. 1997 [citado em: 26 de março de 2023]; 26(1):220-3. Disponível em: https://pubmed.ncbi.nlm.nih.gov/9126523/. Doi: 10.1093/ije/26.1.220. PMID: 9126523.

29. Barros AJ, Hirakata VN. Alternatives for logistics regression in cross-sectional studies: an empirical comparison of models that directly estimate prevalence ratio. BMC Medical Research Methodology [Internet]. 2003 [citado em: 29 de março de 2023]; 3(21):1-13. Disponível em: https://pubmed.ncbi.nlm.nih.gov/14567763/. Doi: 10.1186/1471-2288-3-21. PMCID: PMC521200. PMID: 14567763.

30. Tornesello ML, Losito S, Benincasa G, et al. Human papillomavirus (HPV) genotypes and HPV16 variants and risk of adenocarcinoma and squamous cell carcinoma of the cervix. Gynecol Oncol [Internet]. 2011 [citado em: 10 de dezembro de 2022]; 121(1):32-42. Disponível em: https://pubmed.ncbi.nlm.nih.gov/21211829/. Doi: 10.1016/j.ygyno.2010.12.005. PMID: 21211829

31. Zuna RE, Tuller E, Wentzensen N, et al. HPV16 variant lineage, clinical stage, and survival in women with invasive cervical cancer. Infect Agents Cancer [Internet]. 2011 [citado em: 23 de abril de 2023]; 6:19. Disponível em: 10.1186/1750-9378-6-19.

32. Rader JS, Tsaih SW, Fullin D, et al. Genetic variations in human papillomavirus and cervical cancer outcomes. Int J Cancer [Internet]. 2019 [citado em: 03 de janeiro de 2023]; 144(9): 2206–2214. Disponível em: https://pubmed.ncbi.nlm.nih.gov/30515767/. Doi:10.1002/ijc.32038. PMID: 30515767; PMCID: PMC6450540.

33. Quinn BA, Deng X, Colton A, et al. Increasing Age Predicts Poor Cervical Cancer Prognosis with Subsequent Effect on Treatment and Overall Survival. Brachytherapy [Internet]. 2019 [citado em: 19 de janeiro de 2023];18(1):29-37. Disponível em: https://pubmed.ncbi.nlm.nih.gov/30361045/. Doi: 10.1016/j.brachy.2018.08.016. PMID: 30361045; PMCID: PMC6338515.

34. Barben J, Kamga AM, Dabakuyo-Yonli TS, et al. Cervical cancer in older women: Does age matter? Maturitas [Internet]. 2022 [citado em: 16 de março de 2022]; 158:40-46. Epub 2021. Disponível em: https://pubmed.ncbi.nlm.nih.gov/35241237/. Doi: 10.1016/j.maturitas.2021.11.011. PMID: 35241237.

35. Knüppel S, STANG A. DAG Program: identifying minimal sufficient adjustment sets. Epidemiology [Internet]. 2010 [citado em: 24 de março de 2023]; 21(1):159. Erratum in: Epidemiology. 2010; 21(3):432. Disponível em: https://pubmed.ncbi.nlm.nih.gov/20010223/. Doi: 10.1097/EDE.0b013e3181c307ce. PMID: 20010223.

36. Sharma C, Deutsch I, Horowitz DP, et al. Patterns of care and treatment outcomes for elderly women with cervical cancer. Cancer [Internet]. 2012 [citado em: 22 de fevereiro de 2022]; 15;118(14):3618-26, ISSN 1097–0142. Epub 2011. Disponível em: https://pubmed.ncbi.nlm.nih.gov/22038773/. Doi: 10.1002/cncr.26589. PMID: 22038773.

37. de Sanjose S, Wheeler CM, Quint WGV, et al. Age-specific occurrence of HPV 16-and HPV 18-related cervical cancer. Cancer Epidemiol Biomarkers Prev [Internet]. 2013 [citado em: 06 de março de 2021]; 1313–1318. Disponível em: https://pubmed.ncbi.nlm.nih.gov/23632816/. Doi: 10.1158/1055-9965.EPI-13-0053. PMID: 23632816; PMCID: PMC4306595.

38. Dale DC. Poor prognosis in elderly patients with cancer: the role of bias and undertreatment. J Support Oncol [Internet]. 2003 [citado em: 20 de janeiro de 2023]; 1(4 Suppl 2):11-7. PMID: 15346995. Disponível em: https://www.ncbi.nlm.nih.gov/pubmed/15346995. PMID: 15346995.

39. Albores-Saavedra J, Gersell D, Gilks B, et al. Terminology of Endocrine Tumors of The Uterine Cervix: Results of A Workshop Sponsored by The College of American Pathologists And The National Cancer Institute. Arch Pathol Lab Med [Internet]. 1997 [citado em: 15 de junho de 2022]; 121(1):34-9. Disponível em: https://pubmed.ncbi.nlm.nih.gov/9111090/. PMID: 9111090.

40. Wang SS, Sherman ME, Silverberg SG, et al. Pathological Characteristics of Cervical Adenocarcinoma in A Multi-Center U.S. Based Study. Gynecol Oncol Internet]. 2006 [citado em: 02 de agosto de 2022]; 103(2):541-6. Disponível em: https://pubmed.ncbi.nlm.nih.gov/16697450/. Doi: 10.1016/j.ygyno.2006.03.049. PMID: 16697450.

41. Galic V, Herzog TJ, Lewin SN, et al. Prognostic Significance of Adenocarcinoma Histology in Women with Cervical Cancer. Gynecol Oncol [Internet]. 2012 [citado em: 05 de junho de 2022]; 125(2):287-291. Disponível em: https://pubmed.ncbi.nlm.nih.gov/22266551/. Doi: 10.1016/j.ygyno.2012.01.012. PMID: 22266551.

42. Kodama J, Seki N, Nakamura K, et al. Prognostic Factors in Pathologic Parametrium-Positive Patients with Stage Ib-IIb Cervical Cancer Treated by Radical Surgery and Adjuvant Therapy. Gynecol Oncol [Internet]. 2007 [citado em: 13 de janeiro de 2022]; 105:757-761. Disponível em: https://pubmed.ncbi.nlm.nih.gov/17433424/. Doi: 10.1016/j.ygyno.2007.02.019. PMID: 17433424.

43. Kasamatsu T, Onda T, Sawada M, et al. Radical Hysterectomy for Figo Stage I–IIb Adenocarcinoma of The Uterine Cervix. Br J Cancer [Internet]. 2009 citado em: 08 de janeiro de 2023]; 100:1400–1405. Disponível em: 10.1038/sj.bjc.6605048.

44. Katanyoo K, Sanguanrungsirikul S, Manusirivithaya S. Comparison of Treatment Outcomes Between Squamous Cell Carcinoma and Adenocarcinoma in Locally Advanced Cervical Cancer. Gynecol Oncol [Internet]. 2012 [citado em: 09 de janeiro de 2023]; 125:292-296. Disponível em: https://pubmed.ncbi.nlm.nih.gov/22293041/. Doi: 10.1016/j.ygyno.2012.01.034.PMID: 22293041.

45. Kang S, Wu J, Li J, et al. Prognostic Significance of Clinicopathological Factors Influencing Overall Survival and EventFree Survival of Patients with Cervical Cancer: A Systematic Review and Meta-Analysis. Med Sci Monit [Internet]. 2022 [citado em: 10 de abril de 2023]; 28:e934588. Disponível em: https://pubmed.ncbi.nlm.nih.gov/35260545/. Doi: 10.12659/MSM.934588. PMID: 35260545; PMCID: PMC8919681.

